# Predicting engagement in working memory training for methamphetamine use disorder: indices of motivation, attention and achievement

**DOI:** 10.1101/2020.12.15.20248232

**Authors:** EF Shorer, SJ Brooks

## Abstract

**Rationale:** Impulsivity and inefficient self-regulation are major deficits in substance use disorder, corresponding to lack of treatment response and low abstinence rates. Prefrontal cortex (PFC) executive functions, such as working memory (WM), enable self-regulation of impulsivity by keeping in mind cognitive strategies for optimal decisions about future action. Training WM may improve self-regulation of impulsivity in various populations, including methamphetamine use disorder (MUD) patients. However, many controversies exist regarding the efficacy of WM training (e.g. near-versus far-transfer effects) and associated improvements to clinical outcomes. Such controversies may relate, as observed in our preliminary study in MUD patients, to variation in WM training engagement.

**Methods:** In this nested examination of n=17 male MUD patient data (age range 20-39 years) from a previously published study, the primary aim was to identify variables that may predict a WM training engagement index (EI), and secondarily to determine if EI could predict clinical improvement in MUD patients. The EI consisted of three domains of WM training: motivation, attention/learning and achievement, and we examined the link between self-report measures of impulsivity, self-regulation, anxiety/depression and executive function and EI.

**Results:** Those with higher rates of attention/learning demonstrated greater improvement in attentional impulsivity, and those with higher rates of achievement had a significant improvement in self-reported anxiety, as well as slower time to complete an executive function task after treatment.

**Discussion:** We suggest the EI is a useful measure to detect level of engagement during WM training in patients with MUD and is predictive of clinical improvements.

## Rationale

Impulsivity and inefficient self-regulation are major deficits in substance use disorder (SUD) (Potvin et al. 2018), underscored by aberrant neural processes for the surveillance of internal and external environment and deliberation on appropriate future actions. Moeller et al defines the phenomenon of impulsivity, or *acting without planning ahead* as: “The predisposition towards rapid, unplanned reactions to internal or external stimuli without regard to the negative consequences of these actions to the impulsive individual”(Moeller et al. 2001). Impulsive thinking affects all individuals to varying degrees, however it is significantly more severe in patients with disorders such as Attention Deficit Hyperactivity Disorder (ADHD)(Castellanos et al. 2006), Schizophrenia(Li et al. 2015), and Substance Use Disorder (SUD)(Bechara and Martin 2004), which often have comorbid dysfunctional impulse control. Methamphetamine use disorder (MUD) is the second most prevalent SUD globally(United Nations Office on Drugs and Crime 2016) and is partly characterized by impulsive behaviours that coincide with low abstinence rates after treatment (Brooks et al. 2016). Higher rates of impulsivity may explain why those with MUD tend to reuse drugs after therapy and sometimes commit crimes and acts of physical and sexual violence [7,8]. Thus, given the rising rates of MUD and related impulse-control behaviours, the development of novel interventions as adjuncts are needed to enhance the efficacy of standard treatments that may enable patients to better plan optimal, pro-social actions. However, it is currently unclear how to measure the efficacy of such novel interventions that aim to alter neural processes corresponding to beneficial clinical outcomes for patients in treatment for MUD.

Neurologically, impulsivity is associated with inefficacy within prefrontal cortical (PFC) circuitry such that “top-down” cognitive control is compromised (Bechara and Martin 2004). The PFC forms part of executive control network, which integrates signals from within corticolimbic circuits (Baars et al. 2013). Consolidation of internal and external information within the PFC allows several bits of information to be held in mind in the absence of continued stimulation. This in turn enables optimal decisions about future outcomes to be made with appropriate insight. As such, structural and functional neural deficits observed in those with MUD (London et al. 2015) may limit memories of previous decisions and consequences, and the processing of new cognitive strategies, for future behaviour learned during treatment, to be held in mind. Such deficiencies are thought to be an endophenotype in impulsive individuals(Khurana et al. 2017), and those who are more predisposed to impulsive thinking are more likely to abuse substances and subsequently experience the neurotoxic damage of the drug, thus worsening their impulsivity. To address this cycle of impulsivity, physicians may be able to improve PFC functioning with novel treatment interventions that complement existing therapies.

Currently there are limited options to treat MUD, and the most effective treatment is predominantly based on Cognitive Behavioural Therapy(CBT) (Courtney and Ray 2014), a psychological ‘talking-cure’ that aids patients in updating cognitions concerning affect and behaviour. However, after CBT and related interventions, relapse typically remains high (Brooks et al. 2016), and is likely due to unremitting cognitive dysfunction and enduring neural deficits (Potvin et al. 2018). Addressing cognitive dysfunction in MUD patients seems crucial to rehabilitating the condition, prompting adjunctive therapies that aim to harness and improve PFC function in relation to self-regulation of impulsivity. One such adjunctive therapy is cognitive training (CT), which attempts to improve efficiency in neurocircuitry by repetition of gradually more demanding tasks (Brooks et al. 2017a). CT that aims to improve working memory (WM) might be particularly effective for MUD, given that WM is a PFC process associated with holding information in mind for a prolonged period, is crucial to decision making and self-regulation [12,13], and is dysfunctional in MUD (Yi et al. 2010). Figure 1 provides a schematic illustration of the influence of WM in substance abuse versus abstinence. WM training (WMT) attempts to widen WM capacity and improve the efficiency of circuits within the PFC to hold cognitive strategies learned during CBT in mind for longer [11]. WMT has been trialled in several populations, showing some efficacy in reducing symptoms of impulsivity in ADHD(Spencer-Smith and Klingberg 2015) and SUD patients(Houben et al. 2011), although there are still controversies as to whether WMT is effective (Melby-Lervåg et al. 2016).

**Fig. 1.**
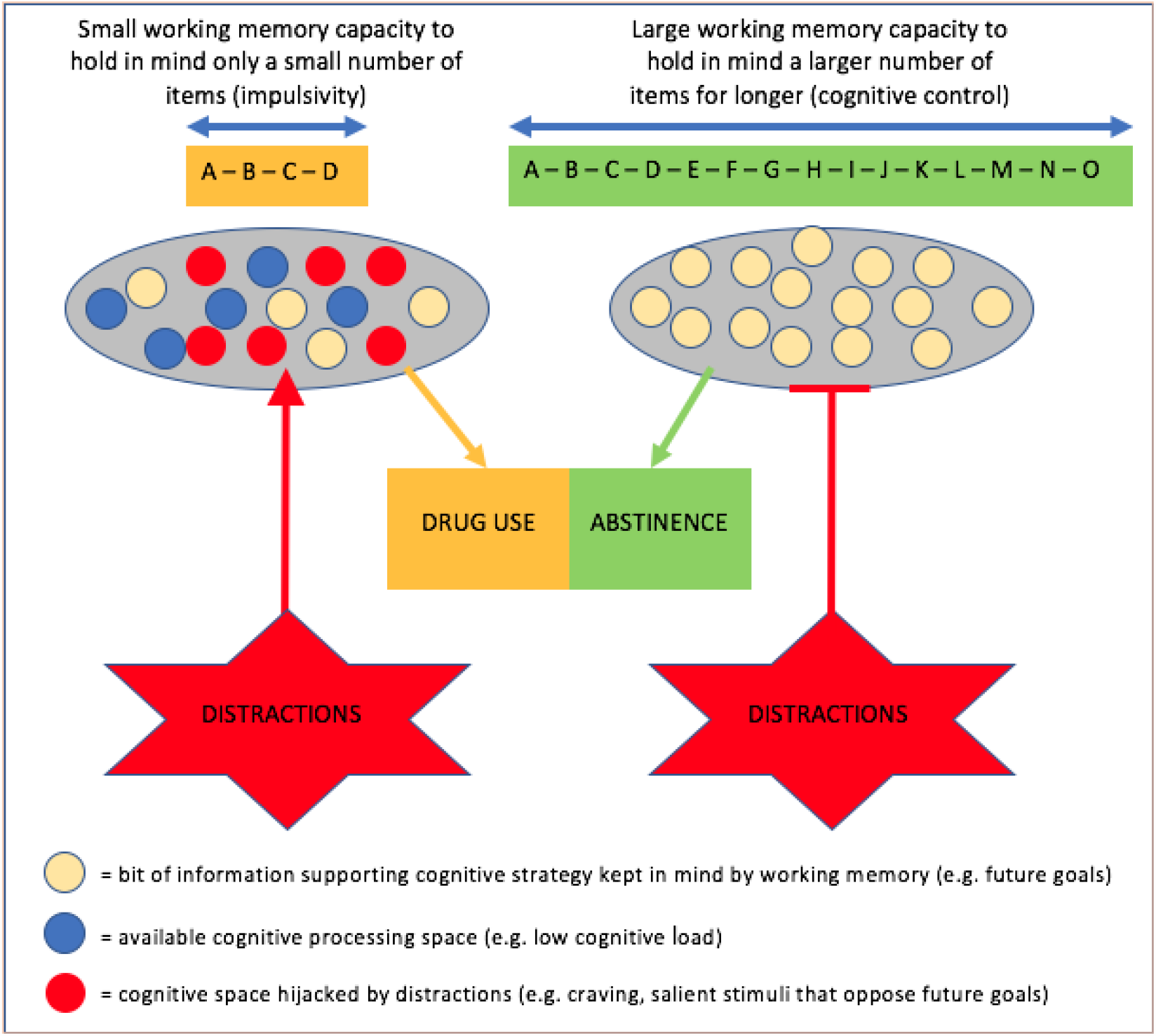
Schematic depicting the relationship between WM capacity and the ability to resist distractions.

In the first preliminary study of WMT in MUD by our group(Brooks et al. 2017c), WMT was given as an adjunct to treatment as usual (TAU), while other MUD patients had TAU only. MUD patients who engaged in WMT reported an improvement in their ability to self-regulate and a reduction in their impulsivity. However, during the study, MUD patients undergoing WMT had differential degrees of engagement, defined as successfully completing training (e.g. gauged by number of missed sessions, highest score, amount of fluctuation between levels of n-back training). The factors underlying this differential engagement are unknown and could help to understand the existing controversies in the field related to efficacy of WMT (e.g. near-versus far-transfer). Moreover, it is not yet shown whether the degree of engagement in WMT is predictive of beneficial clinical outcomes. Implementing WMT in MUD patients who are unlikely to effectively engage in the intervention (e.g. due to severe neurocognitive deficits that first need addressing with standard treatment) constitutes a misuse of both of time and effort on behalf of clinicians and patients. Providing clinicians with a tool to predict which MUD patients are likely to engage in WMT will enable personalized treatment intervention for MUD rehabilitation.

Against this background, the primary aim of this research is to investigate a checklist of factors that predict WMT engagement in the cohort of MUD patients previously studied by Brooks et al (Brooks et al. 2017c). The secondary aim is to determine whether degree of engagement correlates with beneficial clinical outcomes. Herein we hypothesize that (1) extent of engagement in WMT will be associated with specific patient factors, and (2) beneficial clinical outcomes after 4 weeks of the training will be predicted by an engagement index score.

## Methods

### Participants and clinical setting

See Fig .2 for CONSORT (Consolidated Standards of Reporting Trials) recruitment diagram. The current nested study examines the sub-set of MUD patients from the Brooks et al(Brooks et al. 2017c) study that engaged in WMT only. In the main study sixty MUD patients (confirmed prior to admittance) were enrolled in the pilot alongside 30 healthy controls (HC) aged between 18 and 50. Of the 60 MUD patients in the original study, 15 received treatment as usual (TAU) and 20 received working memory training cognitive training (CT) together with TAU, the remaining were excluded for reasons described in the CONSORT diagram. In this nested study only 17 CT patients were included in the final analysis, (*n*=3 did not answer all the baseline questionnaires and hence correlations requiring baseline impulsivity measures could not be performed). All MUD patients enrolled in the study were recruited from an in-patient psychiatric facility and were abstinent from drug use for a minimum of 2 weeks. The treatment program is described in more detail in the previous publication [19] and ran across 8 weeks (2 week abstinent/acclimatisation period; 4 weeks of core treatment; 2 weeks preparation for returning to normal life). The mean duration of MUD exposure prior to admittance and before the final analysis was 9.5 years (*s*.*d*. = 3.65).

**Fig. 2.**
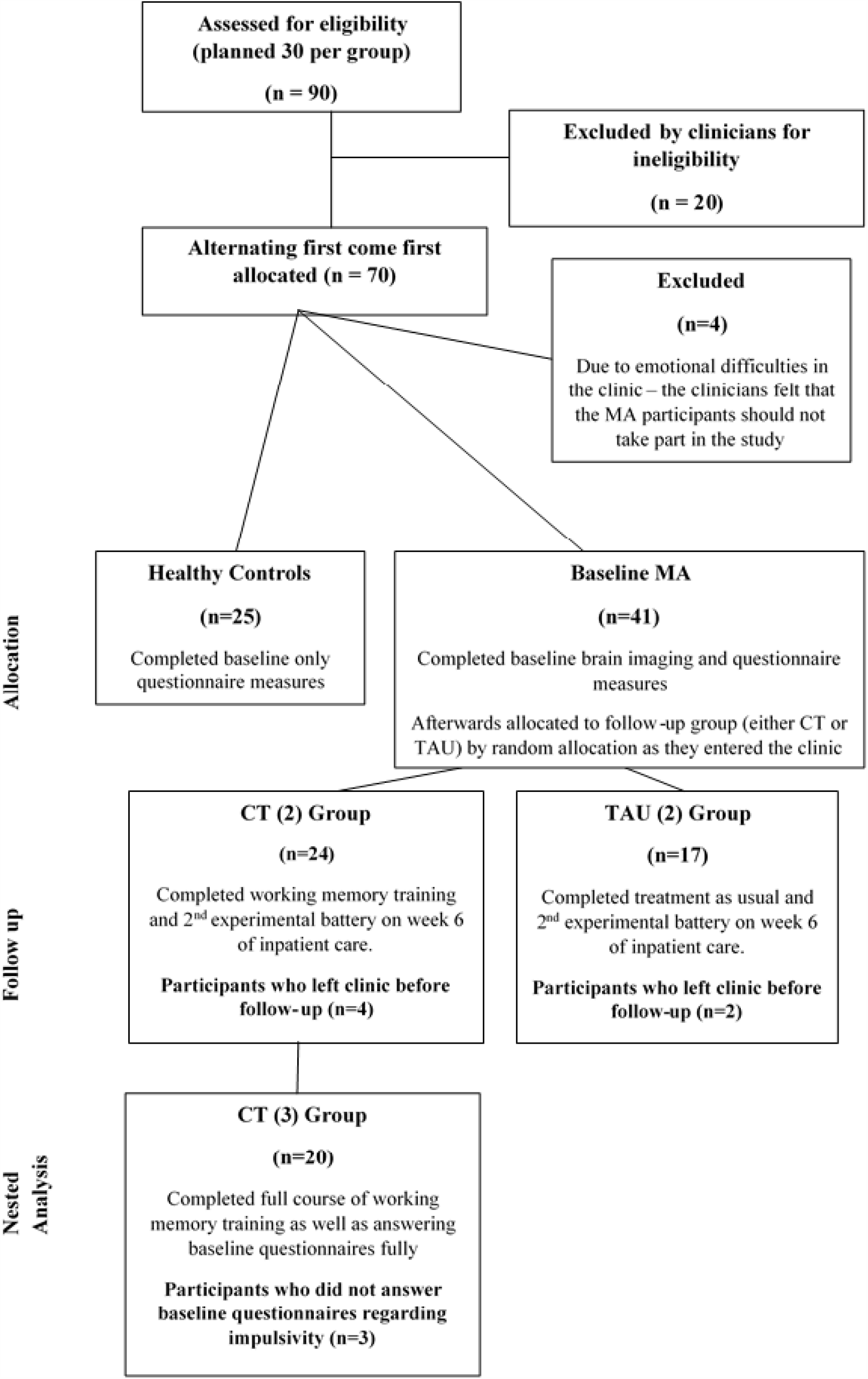
CONSORT diagram demonstrating participant selection for nested analysis.

### N-Back Working Memory Training task

Participants in the original study [18] performed a modified version of the N-Back computer-based WM task designed by Kirchner(Kirchner 1958). During the N-Back task a sequence of letters appear on a computer screen one at a time, each letter for approximately one second. Participants engage their WM as they are required to hold this sequence in mind while simultaneously recognizing whether the letter currently on screen is identical to a previous letter. In the first level “0-back”, the participants simply press the space bar on the keyboard when the letter “X” appears. In the next level, “1-back”, participants press if the letter displayed was identical to the letter that appeared on screen immediately before it. The following levels “2-back” and “3-back” required participants to press the spacebar if the letter was identical to the letter displayed on the screen 2 instances before, and 3 instances before, respectively. Higher levels of N-Back increase WM demand. Participants could voluntarily move up a level upon achieving 80% or more on the current level, and during this study were capped at 3-back, to avoid disappointment if patients were not able to progress to higher levels. Peripheral mosaics were added to the traditional task as a modification intended to mimic real life peripheral distractions.

Accuracy was calculated by the following formula proposed by Miller et al. (Miller et al. 2017):

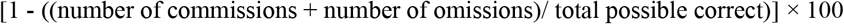

Commissions refer to instances where participants identified an incorrect letter (i.e. pressing to a non-target), and omissions refer to instances where participants fail to identify the correct letter (i.e. failing to press to a target).

### Defining engagement in WM training

We considered engagement in WMT to consist of three aspects, namely: *intrinsic motivation, attention and learning*, and *achievement*. Thus, if a participant demonstrated a motivated performance, attended to the task and demonstrated improvements during the N-back task they were considered to have engaged in WM training. Below we provide further detail on the definition of these three components of WM engagement.

### Intrinsic motivation

Gard et al(Gard et al. 2009) suggest that motivation is linked to evaluation of saliency of stimuli and is moderated by WM. As such, goal-seeking behaviour, e.g. improving one’s performance on a progressively difficult WM task may stimulate saliency regions of the brain, such as the mesolimbic pathway (Berridge and Kringelbach 2008), and is linked to motivation to continue with the task (e.g. a lower number of missed sessions). Thus, perhaps MUD patients with significant WM deficits may not receive enough saliency during the task, and consequently have reduced motivation to learn and/or to continue with the task. Therefore, we suggest, in line with others in the field of WM training (Zhao et al. 2017) that intrinsic motivation is a vital component of engagement in WMT for MUD patients.

### Attention & Learning

Chronic use of methamphetamine damages the prefrontal cortex and consequently circuits that are responsible for governing attention and WM(Scott et al. 2007). McKetin et al(McKetin and Mattick 1998) identified that severity of dependence to methamphetamine was the greatest predictor of attention deficits in MUD patients. Such deficits in attention may reflect an inability to adequately engage in training.

### Achievement

Neuro-circuitry governing WM suggests potential plasticity and learning proportional to the degree of WMT patients receive (Jaeggi et al. 2008). However, it remains unknown whether this axiom is true with regards to MUD patients as methamphetamine significantly damages PFC systems (Scott et al. 2007). Therefore, if MUD patients demonstrate a significant ability to improve during the N-back task, it may suggest that mechanisms underlying neuroplasticity determine whether a patient is likely to engage in the WMT programme.

### The WMT Engagement Index – a unitary measure of performance for each participant

As engagement was not directly studied in the original analysis, we make use of post-hoc measures to quantify each defined aspect of engagement as described above. The WMT Engagement Index (EI) proposed here is a novel measure that incorporates the above aspects of engagement in WMT for MUD patients involved in the main study upon which this nested study is based. The EI refers to a) *motivation*, b) *attention and learning* and c) *achievement as 3* domains of WMT engagement. Each domain will be scored as either a 1 or 0 (i.e. having engaged or not), giving a maximum possible score of 3 for all CT patients involved in this nested study. The criteria below were used to determine whether a 1 or 0 engagement score was given for each domain.

### Motivation (referring to the easier N-back sessions on 0,1& 2 back)

N-Back levels that are traditionally regarded as ‘low-load’ (0, 1, & 2) were required to familiarize participants with the computer-based format during the WM training of the original study. Whereas, the higher level (3-Back) appeared difficult for MUD patients to complete on first trial, due to a wide range of accuracy scores after successful completion of prior levels (see [18]). Some patients had the motivation to move up the N-back levels to 3-back in fewer sessions, while others remained on the low levels for more sessions. As such, spending more than 25% of the WMT (5 sessions) on the introductory levels was deemed an arbitrary threshold for non-engagement on this aspect of the EI. Thus, participants in this study were given the following score on this domain:

0: >5 sessions on 0,1&2 back

1: <=5 sessions on 0,1&2 back

### Attention & Learning

This domain assumes that learning follows the trend suggested by Wright et al(Wright 1936). Wright suggests that the final stages of learning involves a plateau phase where rate of improvement stabilizes. If participants were at the plateau of learning in the WMT task (e.g. scores were higher than baseline and became consistent) then it could be said they have engaged in training and shown improvement over time. This plateau phase is marked by minimally changing scores at the terminal end of training and is reflected by the standard deviation of the last 25% of the training (5 sessions). High WM training accuracy standard deviation at the end of the training program suggests that performance fluctuated to a large degree and the plateau phase of learning had not begun. Smaller standard deviations of the accuracy scores, however, could reflect that the participant had reached Wright’s plateau phase, and therefore had engaged. On this basis, participants were given the following scores:

0: >5 standard deviation on last 5 trials

1: <=5 standard deviation on last 5 trials

### Achievement

If participants regularly achieved high accuracy scores (e.g. 80% or higher) in the most difficult level of training during WMT - 3-Back - it would suggest a sufficient level of achievement. The threshold of 80% was chosen as this value has been shown to represent significant adeptness during WMT (Olesen et al. 2014). Peak performance was examined by evaluating how many sessions on 3-Back the participant achieved an accuracy score of 80% or greater. As such, participants were given the following scores:

0: < 2 scores over 80% on 3-back

1: >= 2 scores over 80% on 3-back

Self-report measures

For the second aim of this nested study, we examined whether WMT engagement index was associated with clinically relevant outcomes, based on self-report measures as per the main study, as follows:

### Visual Analogue Scale (VAS) for mood, desire for drug and self-control(Reips and Funke 2008)

This psychometric measure comprises of 3 distinct scores for subjective feelings of mood, desire for drug and self-control. A horizontal line is used to indicate a spectrum, the left side of the line indicating low levels of happiness, low desire for the drug and low feelings of self-control. The right side of the line indicates the opposite extreme. Participants are asked to indicate on the spectrum where they currently feel. This mark is converted to a percentage for statistical analysis.

### Trail Making Test (TMT)(Tombaugh 2004)

The Trail Making Test is not a self-report measure but a brief neurocognitive task designed to assess visual searching and scanning speed, mental flexibility and executive functions (including working memory). It consists of two subtests: TMT-A and TMT-B. The TMT-A test involves timing how quickly participants can draw lines to connect 25 numbers in sequence. TMT-B is a similar timed test, however the participant alternates between numbers and letters (e.g. 1, A, 2, B, etc.). Times for the TMT-B are generally slower due to the increased complexity of this task. Furthermore, the difference between the times taken to complete each test (TMT B time – TMT A time) has been used as a marker of executive function(Journal et al. 1996). Age and education can partially account for 35-40% of the variance in this test(Tombaugh 2004),(Journal et al. 1996).

### Hospital Anxiety and Depression Scale (HADS)(Snaith and Zigmond 1983)

This test requires patients to answer 7 questions related to anxiety and 7 questions related to depression. Each question is answered on a 4-point scale. The maximum score attainable for anxiety and depression respectively is

21. Scores of 0-7 are considered normal, scores of 8-10 and considered borderline, and scores of 11-21 are considered indicative of a potential diagnosis of anxiety and/or depression. A review of the scale suggested that both the anxiety and depression subscales have a specificity and sensitivity of 0.8 for their respective conditions, and was well evaluated at detecting the severity of anxiety and depression(Bjelland et al. 2002).

### Barratt Impulsiveness Scale (BIS)(Patton et al. 1995)

The questionnaire is also based on a 4-point scale with 30 questions related to several aspects of impulsivity. These aspects are divided into 6 primary factors (Attention, Motor, Self-control, Cognitive-complexity, Perseverance, Cognitive-instability) and 3 second order factors (Attentional Impulsiveness, Motor Impulsiveness, Non-planning Impulsiveness). Each second order factor is regarded as a combination of 2 of the primary factors. In a review of the BIS by Stanford et al (Stanford et al. 2009), it was found that the second order factors of Non-planning best reflected WM capacity. Also, of note was that the second order factor of Motor Impulsiveness was predicted by TMT scores and was related to WM capacity. The total BIS score has shown to be of utility in recognizing various kinds of impulsive behaviour in patients.

### Self-Regulation Questionnaire (SRQ)(Carey et al. 2004)

This questionnaire encompasses 63 questions based on a 5-point scale that are designed to gauge an individual’s ability to self-regulate. The questions are grouped into 7 broad skills: ability to receive relevant information, evaluate information, trigger change, search for different options, formulate a plan, implement the plan, and further assess it. Although the questions can be distributed into the 7 factor subscales, there is little proven evidence for its validity as independent measures.

### Baseline Demographic Variables

The following patient information was gathered at baseline to be used in the subsequent exploratory data analyses: (a) patient age; (b) level of education (evaluated as either having completed high school or not) (c) dietary status (evaluated as either poor or balanced) (d) number of dependencies (e) years of drug abuse and (f) weeks abstinent.

### Evaluating Engagement Index against clinical outcomes

Secondary outcome measures were used to validate the Engagement Index against clinical measures of improvement. These secondary measures were referred to as “Delta scores” as they reflect the percentage improvement or percentage deterioration in the various self-report measures between baseline and 4-week follow up after WMT. Each Delta score was calculated by using the following formula:

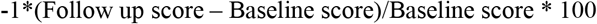

This formula gives the change in score as a percentage. If there was an improvement the percentage was positive, while if the measure showed deterioration the value was negative. These scores were calculated for the following self-report measures and their subscales: Trail Making Test (A, B, Difference), HADS, BIS & SRQ.

### Statistical analyses

#### Assumptions

Data points with a Z-Score of 2.5 or greater were considered outliers and removed from the investigation (considered null). Three participants did not complete questions for the baseline measures and thus these values remained null. All null values were excluded from any tests of correlation by performing pairwise exclusion methods. Shapiro-Wilk tests for normality were conducted on all the variables investigated. Levene’s test was performed on the normal variables to test the assumption of heteroscedasticity. Kendal’s Tau correlations were conducted if one or more of the variables were non-normal or displayed homoscedasticity. Pearson correlations were performed on the normally distributed data if the assumption of heteroscedasticity was met. These correlations were used to address the aims and hypotheses of this investigations.

#### Statistical Modeling

Classical linear regression models could not be used to predict EI as the scores were non-normally distributed and the dependent variable was ordinal (0-3). Instead, a multinomial logarithmic regression (logit) model was designed to calculate the odds ratios for each EI score. Only 1 baseline variable could be carried forward in the model based on the rule of thumb stating predictive models should have approximately 10-15 samples per predictor (Field 2005). The baseline variable that would be carried forth into the logit model was that which gave the highest correlation to the Engagement Index in the exploratory data analyses. Issues of multicollinearity would not be present with a solitary predictor.

## Results

### Evaluation of the EI factors

Delta scores of the self-report measures were correlated against each of the EI domains to observe trends between EI scores and improvement and deterioration with respect to executive function, impulsivity, self-regulation, anxiety and depression. The full table of correlations is shown in Table 1. There were no significant correlations between any of the clinical measures and the motivation score (first domain of EI). Attention and learning (second domain of EI) was significantly correlated to improvement in 2^nd^ Order Attentional BIS (*r* =.487, *τ* < .05). On average, participants who engaged in the trial based on the attention and learning domain showed 24.48% improvements in attentional impulsivity as opposed to a 4.91% deterioration for those who did not engage (Figure 4). WM achievement (third domain of EI) was significantly correlated to improvements in anxiety measured by the HADSa (*r* =.451, *τ* < .05), and in TMT time difference (*r* =-.609, *τ* < .05). On average, those who engaged based on the WM achievement criteria had a 24.41% improvement in self-reported anxiety and were 99.51% slower in their TMT time difference. Conversely, those who did not engage in this domain were 8.74% more anxious, but 16.88% quicker in their TMT time difference. Global EI scores (adding together all three domain scores), were not significantly correlated to any of the secondary outcome measures.

**Table 1:**
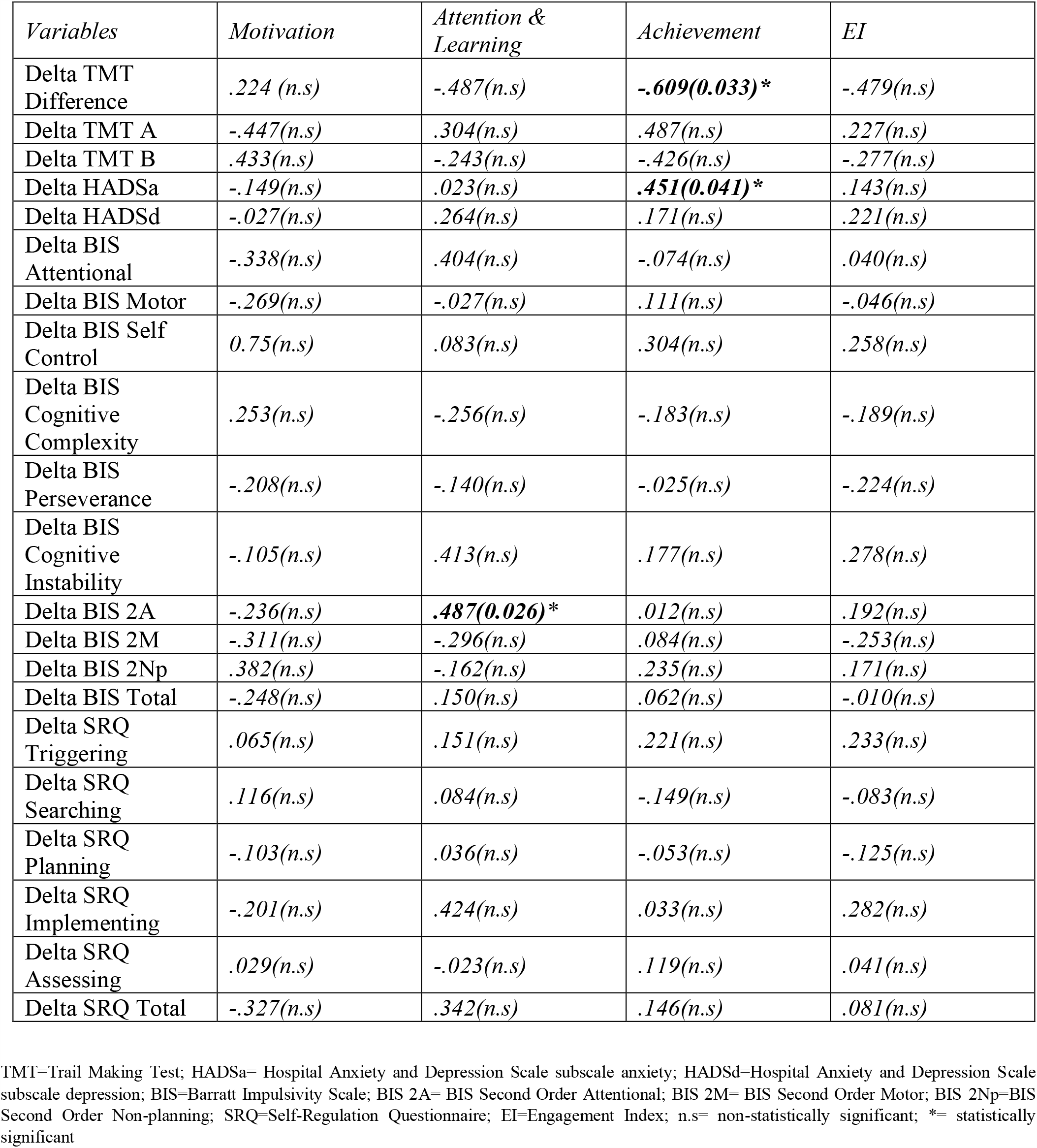
Correlations between Engagement Indices and Clinical Outcome Measures in patients with methamphetamine use disorder (n=17)

**Fig. 3.**
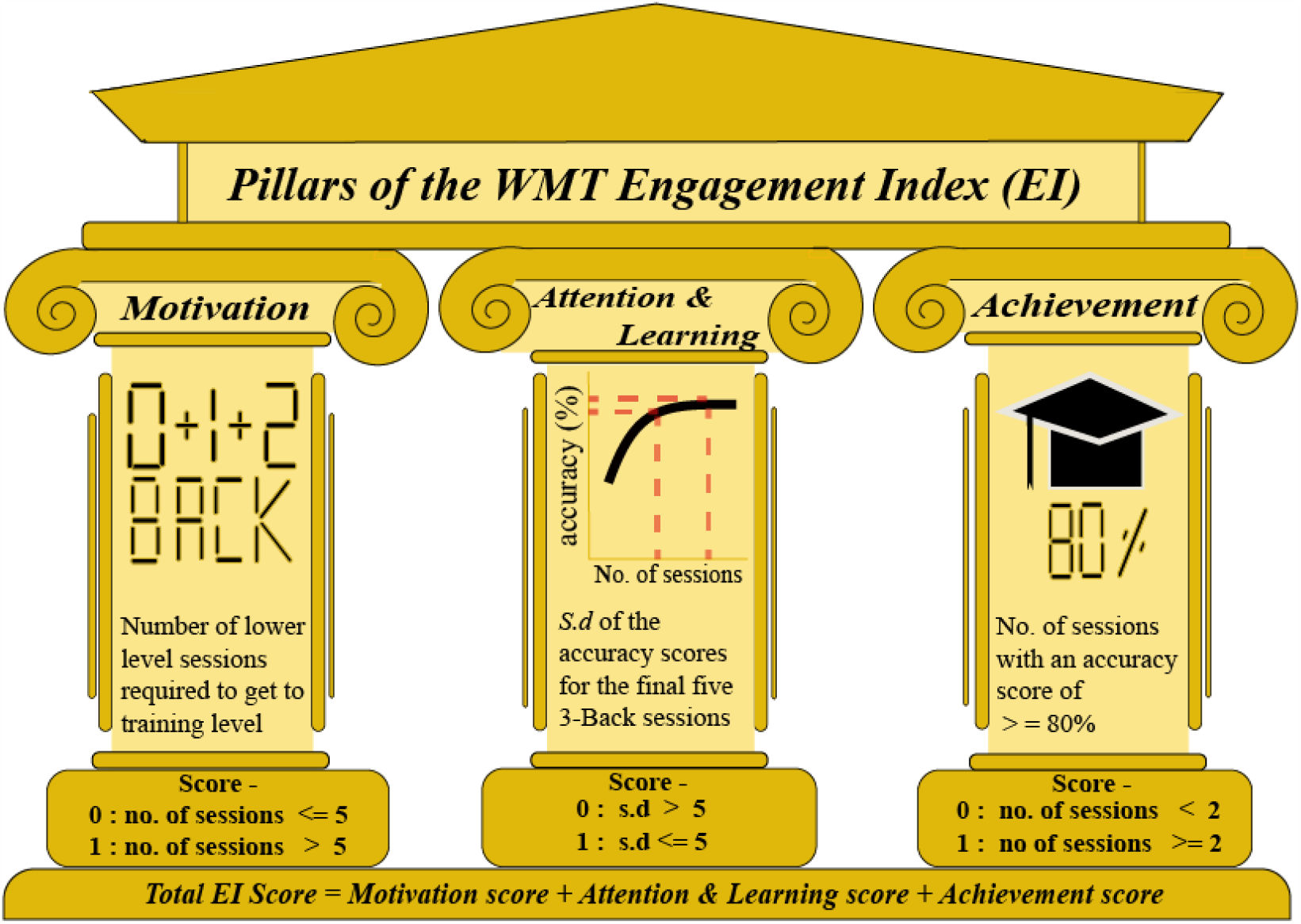
Pillars of the WMT EI.

**Fig. 4.**
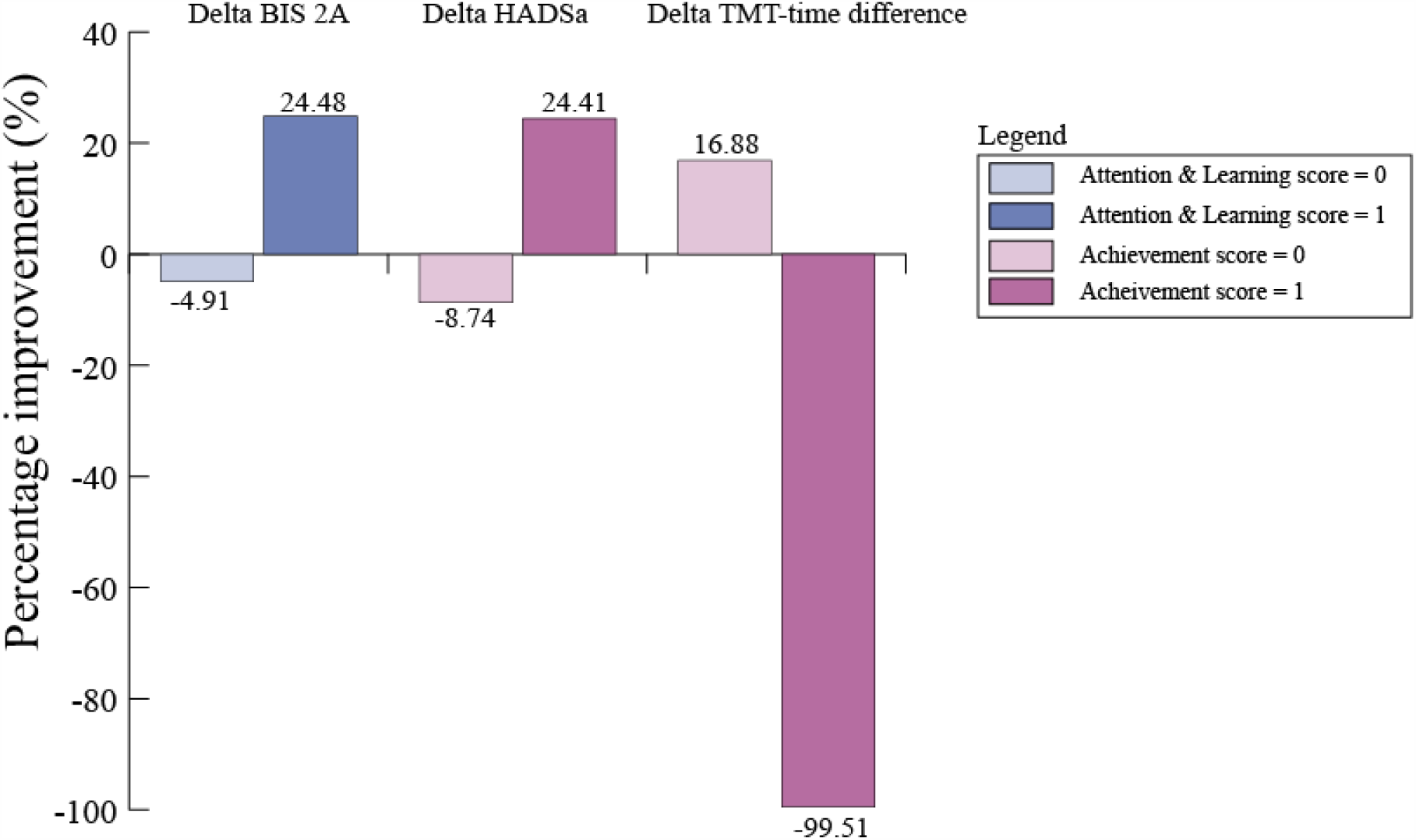
Average percentage improvement in the relevant clinical outcomes of those who engaged versus those who did not. BIS 2A=Barratt Impulsiveness Scale 2^nd^ Order Attentional subscale; HADSa=Hospital Anxiety and Depression Scale anxiety subscale; TMT=Trail Making Test.

### Predictive Model

The baseline clinical measures that had a significant relationship to the EI score (e.g. comprising of scores from the 3 domains) were: dietary status (r = .629; τ = .016), TMT B time (r = -.485; τ =.0.044), TMT B errors (r = -.654; τ = 0.017) and TMT time difference (r = -.628; τ = 0.009). Only one factor could be carried forward into the model as statistically robust models require 10-15 variables per predictor(Field 2005). Therefore, it was decided that TMT time difference would be most appropriate due to its significance level. Five participants had missing information for their TMT times, and thus 12 participants were used in the model design. Figure 5 below ranks the TMT time differences ascendingly adjacent to the corresponding EI score per participant.

**Fig. 5.**
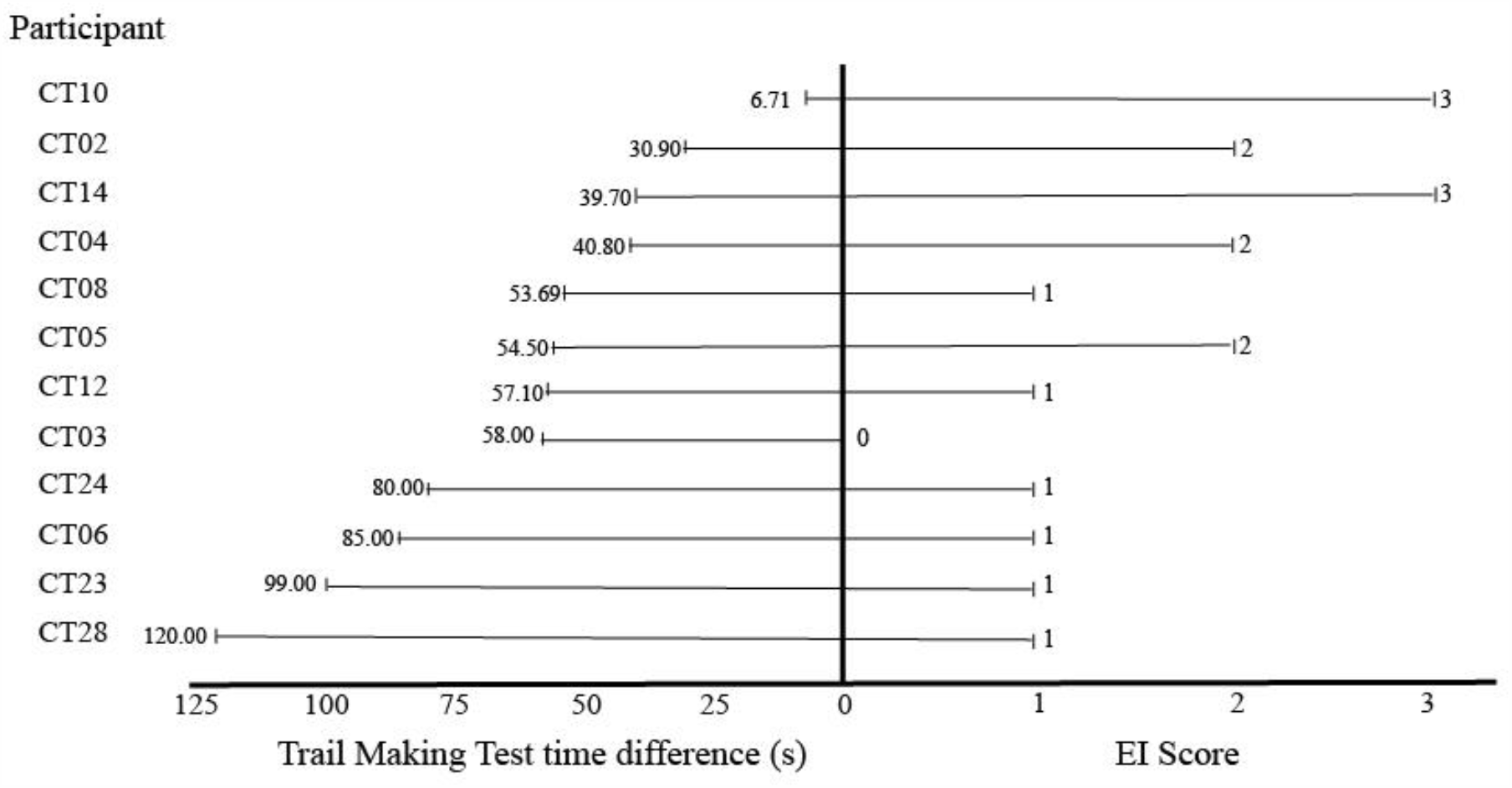
Forrest plot comparing the TMT time difference (TMT-B time – TMT-A time) and EI score per participant. TMT = Trail Making Test; EI = Engagement Index.

## Discussion

We conducted a nested study within an already published study (Brooks et al. 2017c) that had demonstrated neural and neuropsychological improvements following working memory training (WMT) in patients with methamphetamine use disorder (MUD). This nested study examined, for the first time, MUD patient engagement in WMT, in an attempt to understand factors that might diminish efficacy of WMT (e.g. near-versus far-transfer effects), and to examine the clinical value of WMT. Measures of WMT engagement in this nested study consisted of three domains, namely: an intrinsic motivation to learn, available attention to learn, and a demonstration of successful learning. Based on the scores of these domains, a global Engagement Index (EI) was created by the authors as a measure to quantify engagement for each participant.

### Engagement predicts improvement in impulsivity and anxiety

Although the EI, a summation of three scores, did not show any association to beneficial clinical outcomes, two individual components of the EI (attention/learning, and achievement) showed clinical relevancy. First, patients who demonstrated higher rates of attention and learning, and thus adhered most to Wright’s learning curve plateau phase during WMT, showed significantly greater improvements in the 2^nd^ Order Attentional subscale of Barratt Impulsivity Scale after 4 weeks (*r* =.487, *τ* < .05). Whereas others have demonstrated no significant far-transfer effects in global intelligence and attention after WMT (Melby-Lervåg et al. 2016), our finding might hint towards far-transfer effects in the domain of improved attentional impulsivity – at least in substance use disorders that demonstrate a plateau in their learning curve (e.g. stabilising of performance) – which is linked to, but not quite the same as global intelligence and attention per se.

Secondly, an ability to score higher on the achievement domain of the EI correlated with a reduction in self-reported anxiety (*r* =.451, *τ* < .05), which may suggest a progressive effect of mastery and self-regulation, particularly with regard to anxiety and arousal, during treatment intervention. The achievement domain also showed an association to lengthening of TMT time difference (i.e. those with high achievement scores had a wider time gap between TMT-A and TMT-B at follow-up compared to their baseline scores) (*r* =-.609, *τ* < .05), with no significant differences in TMT-A or TMT-B error rate/accuracy. This result is unusual as typically a lengthening TMT-B time suggests a deterioration in visual scanning speed and executive function. However, it seems unlikely that the intervention could have had a deleterious effect on these faculties. We posit that this finding is incidental or suggest speculatively that those who engaged in WMT may be better able to deliberate and reflect on their TMT performance at follow up, relative to how they would have performed at baseline, thus slowing their performance. This correlation suggests engagement in the WMT program is associated with better far-transfer outcomes (e.g. not related to the actual WMT) in terms of reducing anxiety and improvement in deliberative, reflective thinking. Future research is needed to validate this result and investigate possible mechanisms behind this association.

### Trail Making Test predicts engagement

We found that the TMT time difference (and not error rate/accuracy) was the best predictor of engagement in WMT. TMT-B times corrected for TMT-A significantly correlated with engagement, and is a measure of “visual search, scanning, speed of processing, mental flexibility and executive function”(Tombaugh 2004). Mental flexibility and speed of processing are skills which appear crucial for N-Back successful engagement, thus those at baseline with characteristics suggestive of good N-back performance are likely to engage based on the EI criteria.

### Assessment of the EI as a measure of engagement and clinical utility

The Engagement Index was used as a tool to measure patient engagement in WMT, consisting of the 3 equally weighted scores based on theoretical constructs which may affect engagement namely: intrinsic motivation, attention and learning and achievement in WMT. The validity of each measure is discussed independently.

### Intrinsic Motivation

Burbridge (Burbridge and Barch 2007) et al identified that WM moderates the interaction between motivation and emotional experience of positive stimuli in schizophrenic patients. Moreover, Gard et al (Gard et al. 2009) proposes that WM deficits may contribute to a lack of pleasure when attaining goals in schizophrenic patients. Diminished pleasure results in positive behaviours that are not being reinforced, leading to lower motivational states. Although the link between motivation and WM has not explicitly been studied in MUD, MUD patients display similar WM deficits to schizophrenic patients(Kalechstein et al. 2010), and thus WM may play a similar role in their moderating emotional experiences of positive stimuli. In this study, however, motivation aspects of engagement were not associated with clinical improvements. This could be due to several reasons: firstly, the measure used to gauge motivation in this nested study (e.g. number of earlier sessions of lower-load N-Back) may not have been an accurate measure of participant motivational status. Secondly, participants may have not generated sufficient positive emotional response from success at the abstract N-Back used here (e.g. based on presentation of random sequential letters), which in itself is not affective in nature, and as such this may have limited the drive to gain further success. This is particularly pertinent given that the link between N-back performance and clinical improvement is not obvious. Thirdly, the assumption that WM moderates the interaction between motivation and emotional experience of positive stimuli may be incorrect. This is plausible as the neurotoxic effects of MUD may have significantly altered these neural pathways. Further studies should make use of alternative measures to investigate a potential relationship between motivation during N-Back training and clinical outcomes.

### Attention & Learning

McKetin et al(McKetin and Mattick 1998), identified that severity of dependence to methamphetamine was the greatest predictor of attention deficits in MUD patients. Such deficits in attention may reflect an inability to adequately learn and improve in WMT. The measure used in this nested study for attention and learning was based on Wright’s learning curve (Wright 1936), and seems to be a valid measure due to its correlation to attentional aspects of impulsivity on the Barratt Impulsivity Scale observed during this study. A major limitation of this measure is that participants may reach a plateau at very low scores, therefore not improving, but “coasting”, and yet would still be considered to have engaged based on this domain.

### Achievement

We employed the WMT achievement element of engagement whereby participants needed to score 80% or above on several N-back training sessions (regardless of load) to demonstrate engagement based on this criterion. The threshold of 80% was chosen as this value has been shown by others to represent adeptness at the task in other WMT studies (Olesen et al. 2014). Increasing one’s level in the N-back task requires one to achieve 80% or more on the current level. This means that patients not only have to attend to the training, but also need to do so at a suitable rate such that they can achieve this percentage and progress to more demanding levels of the N-Back task. Adequate engagement in the WMT program would entail an ability to progress past the earlier N-Back levels such that they can reach the WM-intensive levels.

### EI score

The EI index considers all three domains as equal components of engagement, which complies to the definition of engagement in this paper. However, if the EI were to be extrapolated to other WMT programs perhaps the attention and learning domain weighted by the WMT achievement domain would be most appropriate, as it would counter for the bias of the plateau phase of the learning curve, whilst simultaneously evaluating success at a trained task. As intrinsic motivation was not linked to any clinical outcome and as such it may be best to exclude this domain when evaluating engagement on future N-Back based WMT programs.

### Study limitations

The selection of participants was a limitation of this study. Firstly, a narrow subset of MUD patients in the Western Cape were included. Only male participants were enrolled in the trial, most of whom were of coloured (South African term for “mixed race”) ethnicity. Although this selection perhaps is indicative of the demographics in the Western Cape, precaution should be taken when extrapolating to other populations. Future research should look to understand whether the determinants and influence of engagement are similar in differing contexts. Secondly, although all participants completed baseline questionnaires there were several missing cases. For example, data was only present for 12 of the 17 participants with respect to baseline TMT times. As TMT difference was the critical dependant variable in the model the missing data affected the statistical strength of the model. Furthermore, performance on the TMT may be acutely biased. If MUD patients are admitted to an in-patient clinic and are asked to perform the TMT immediately, they are likely to perform worse than if they only completed the test after 2 weeks of abstinence. Therefore, this model is limited in that it cannot predict engagement in WMT on admission. Thirdly, the limited sample size meant that a solitary predictive variable could only be used in the model. If the sample size were larger several of the excluded baseline variables which correlated to EI could be used to build a more robust model that explained a greater proportion of the EI variance. Moreover, if a variable that is not biased by acute drug use were added to the model, it would be able to predict engagement on admission. Additionally, we determine the influence of engagement by examining the clinical outcomes at the end of the 4 weeks of training. During this period participants received treatment as usual, which in the original study conducted by Brooks et al, was accounted for by using a control group. Herein we acknowledge that treatment as usual may be a confounder as we simply examine the associations between engagement and outcome. Furthermore, we cannot speculate at the influence of engagement on future clinical outcomes and far transfer effects.

### Implications and directions for future research

The WMT EI derived in this preliminary, exploratory study, is the first measure of engagement in a WMT intervention for MUD described in the literature. Due to the novelty of the EI and the limitations regarding the predictive model, it suggested that study-specific measures of engagement should be used together with principles of the EI to adequately evaluate engagement in future WMT studies. Further research should be dedicated to understanding the determinants of engagement, such that practitioners can optimise the use of WMT on candidates likely to benefit. This research ought to be conducted in various populations, both in healthy and disease contexts, to increase the validity of the identified determinants of engagement. A major benefit of the EI is that it is not specific to the subset of MUD patients as each domain of the EI uses metrics which tracks general engagement in a learning program, regardless of the participant status. Thus, EI can be used in a variety of WMT programs irrespective of the disease contexts.

Critically, engagement in WMT performance does not necessarily translate into far transfer effects and definitive WM improvement. Thus, one must take precaution in interpreting the EI measures. Engagement reflects only the propensity to reliably learn and improve in the N-back task. In other words, participants may engage well in the training and have appreciable EI scores, yet not demonstrate other WM improvements. However, it is believed that those with low EI scores are not likely to show significant WM improvement, as their engagement in the task is low. Therefore, there may be utility in using the EI as a metric to screen participants and predict their level of engagement in the trial. If future research can correlate high EI scores with structural and functional changes in neurological substrates of WM, then perhaps the EI could also be used as a reliable indicator of improved WM.

## Conclusions

In this preliminary, exploratory study, we demonstrate that aspects of engagement in WMT predicted reduction of self-reported anxiety and improved impulsivity in a cohort of MUD patients. Furthermore, we show that the TMT time difference between baseline and 4-weeks (e.g. slower over treatment) predicted engagement in WMT. Engagement was measured using an Engagement Index (EI) score, a novel measure based on three criteria namely: intrinsic motivation, attention and learning, and achievement in WMT. In addition, we have attempted to create a predictive model for WMT engagement in MUD patients, such that clinicians have greater insight into which participants are likely to engage in WMT based on initial neuropsychological testing. Further research is needed to critically appraise and subsequently modify the design of an EI such that it can be adopted as a robust measure of engagement in WMT in various contexts.

## Data Availability

The datasets used and/or analysed during the current study are available from the corresponding author on reasonable request.

## List of abbreviations

BIS: Barratt Impulsiveness Scale
CT: cognitive training
EI: Engagement Index
HADSa: Hospital and Anxiety and Depression Scale (anxiety)
HADSd: Hospital and Anxiety and Depression Scale (depression)
MUD: Methamphetamine Use Disorder
SRQ: Self Regulation Questionnaire
SUD: Substance Use Disorder
TAU: treatment as usual
TMT: Trail Making Test
WM: working memory
WMT: working memory training

